# Evaluating the burden of COVID-19 on hospital resources in Bahia, Brazil: A modelling-based analysis of 14.8 million individuals

**DOI:** 10.1101/2020.05.25.20105213

**Authors:** Juliane F. Oliveira, Daniel C. P. Jorge, Rafael V. Veiga, Moreno S. Rodrigues, Matheus F. Torquato, Nívea B. da Silva, Rosemeire L. Fiaconne, Caio P. Castro, Aureliano S. S. Paiva, Luciana L. Cardim, Alan A. S. Amad, Ernesto A. B. F. Lima, Diego S. Souza, Suani T. R. Pinho, Pablo I. P. Ramos, Roberto F. S. Andrade, Task-force Rede CoVida Modelling

## Abstract

Here we present a general compartment model with a time-varying transmission rate to describe the dynamics of the COVID-19 epidemic, parameterized with the demographics of Bahia, a state in northeast Brazil. The dynamics of the model are influenced by the number of asymptomatic cases, hospitalization requirements and mortality due to the disease. A locally-informed model was determined using actual hospitalization records. Together with cases and casualty data, optimized estimates for model parameters were obtained within a metaheuristic framework based on Particle Swarm Optimization. Our strategy is supported by a statistical sensitivity analysis on the model parameters, adequate to properly account for the simulated scenarios. First, we evaluated the effect of previously enforced interventions on the transmission rate. Then, we studied its effects on the number of deaths as well as hospitalization requirements, considering the state as a whole. Special attention is given to the impact of asymptomatic individuals on the dynamic of COVID-19 transmission, as these were estimated to contribute to a 68% increase in the basic reproductive number. Finally, we delineated scenarios that can set guides to protect the health care system, particularly by keeping demand below total bed occupancy. Our results underscore the challenges related to maintaining a fully capable health infrastructure during the ongoing COVID-19 pandemic, specially in a low-resource setting such as the one focused in this work. The evidences produced by our modelling-based analysis show that decreasing the transmission rate is paramount to success in maintaining health resources availability, but that current local efforts, leading to a 38% decrease in the transmission rate, are still insufficient to prevent its collapse at peak demand. Carefully planned and timely applied interventions, that result in stark decreases in transmission rate, were found to be the most effective in preventing hospital bed shortages for the longest periods.

## Introduction

In December, 2019, clusters of a respiratory disease attributed to a potentially novel coronavirus were identified in the city of Wuhan, province of Hubei, China. This hypothesis was rapidly confirmed and the virus was named as severe acute respiratory syndrome coronavirus 2 (SARS-CoV-2), causal agent of coronavirus disease 2019 (COVID-19). This novel coronavirus rapidly spread across Asia, Europe and others continents, acquiring pandemic scale, as determined by the World Health Organization, in March 11^th^ 2020. As of May, 2020, all parts of the world are, with varying degrees, impacted by the COVID-19 epidemic. More than 4.4 million cases and 300,000 deaths due to the disease have been reported worldwide^1^, in what can be construed as the worst pandemic since the Spanish flu (1918-20).

A prominent feature of the current pandemic is the high person-to-person transmissibility of the virus, with a basic reproduction number estimated at 2.2-2.5 in Wuhan^2,3^. Also worrying are the severity of clinical complications and the lack of either vaccines or effective drugs to, respectively, prevent disease and accelerate the recovery of infected patients. As a consequence, the only effective mechanism available at the moment to dampen virus spread are based on non-pharmaceutical interventions (NPI), and population adherence thereof. Amongst these are included social isolation and distancing, quarantine, travel restrictions as well as changes in individuals behaviors, such as broad use of face masks and heightened preoccupation to hygiene^4^.

The case fatality rate (CFR) of COVID-19 varies significantly between countries, reflecting heterogeneity at various levels including those of testing rates, demography, health services’ preparedness and coverage, as well as the range of NPI implemented, all of which ultimately impact in the number of active cases and in the need for hospitalization of the more severe cases. At the time of writing, the worldwide CFR was estimated at 6.9%, with France (19.2%), Belgium (16.4%), United Kingdom (14.4%), Italy (14.0%) and Hungary (12.9%) having the currently highest CFR in the world. In South America, Brazil is the country with the leading CFR at 7.0%, followed by Argentine (4.8%) and Bolivia (4.5%). However, historical inequalities in the life conditions, economy, and access to health services in these countries make the COVID-19 pandemic particularly tragic in poorer settings.

The elevated CFR of some countries have been partly a consequence of the sudden increase in demand for hospitalization, leading to collapsed health systems due to insufficient physical and healthcare resources. This has a particularly high impact on countries with limited healthcare infrastructure, such as those in Latin America. Brazil, the largest country in the region, provides a cautionary example of the profound impacts of COVID-19 in health systems.

Brazil presented its first confirmed case of COVID-19 on February 26^th^, 2020 in the state of São Paulo, and nationwide community transmission was declared on March 20^th^. At the time of writing, the number of confirmed COVID-19 cases exceeds 200,000 nationally.

Bahia, located in northeast Brazil, has a population of 14.8 million, comprises 417 municipalities and a territorial extension of 567,295 km^2^, which is comparable to that of France. In spite of the economic development (holding the sixth highest gross domestic product among all Brazilian states), the state of Bahia presents marked intra-regional disparities in the access to health, with hospitals and health care investments unequally distributed in the state^5^. Thus, Bahia, Brazil is a representative example of how COVID-19 impacts health resources in developing countries, and what type of measures can be implemented to mitigate its damaging consequences in similar contexts.

Mathematical models are proving instrumental in studying the current COVID-19 pandemic^6^, as well as in driving governmental actions. A hallmark of the latter was the radical shift in the initial “herd immunity” strategy of the United Kingdom, as models produced by the Imperial College London projected a death toll of 500,000 in order to reach this scenario^7^. Substantial insights on the dynamics of disease spread can be gained by using compartmentalized models such as the 3-compartment SIR (susceptible-infected-recovered)^8^. Models that build on these principles have flourished on the recent literature and extend the number of compartments in order to study other key aspects of COVID-19, including the role of asymptomatic transmission^9,10^, social distancing and quarantine strategies^3,11–14^, as well as postepidemic scenarios, e.g. the probability of novel outbreaks^15,16^. The need for hospitalization under various conditions have also been evaluated using mathematical models^17–19^.

In this work we further explore hospitalization needs in a low-resource country during the COVID-19 pandemic, with particular emphasis given to clinical (or ward) and intensive care units (ICU) hospital beds requirements. Particularly, we describe an 8-compartment model with time-varying disease transmissibility that considers asymptomatic transmission, hospitalization of severe cases (requiring clinical/ICU beds), and mortality. The parameters of this model are partly locally-informed with data from hospitals dedicated to treating COVID-19 patients in the region, and partly calibrated against data (cases, deaths) provided by local health authorities, with optimal parameters identified using a particle swarm optimization metaheuristic. This model was applied to study the ongoing COVID-19 outbreak in the state of Bahia, Brazil, an example of low-resource setting with important inequalities in health access, but is extensible and directly applicable to other regions, with the potential to help set targets that function as guides to analyse the evolution of the COVID-19 pandemic, as well as inform the extent of governmental measures needed.

## Methods

### Data sources and case definition

The models produced in this study were informed by data from multiple sources: The daily series of the cumulative confirmed COVID-19 cases and the daily mortality series for the state of Bahia, its capital Salvador and the remaining inland cities were obtained from the Secretary of Health of the State of Bahia (SESAB). Throughout our analyses we consider separately the capital (which concentrate the number of cases in the state) and inland cities. Local health authorities use the following definition of COVID-19 case, based on two criteria: (i) **clinical/epidemiological**, namely a case of suspected flu-like syndrome or severe acute respiratory syndrome (SARS) who had contact with a laboratory-confirmed COVID-19 case in the last 7 days prior to symptoms onset; or (ii) **clinical/laboratory**, a suspected case of flu-like syndrome or SARS with a positive SARS-CoV-2 serology (IgM and/or IgG) or real-time PCR result.

Additionally, we also obtained the state-level daily hospital bed occupancy for both clinical and ICU beds. At the time of writing, the state has a total of 888 hospital beds available for specifically treatment of COVID-19 patients, of which 466 comprise clinical hospitalization and 422 beds for ICUs. This data was not available at the municipality-level; rather, due to the Brazilian administrative division of health regions, hospital bed occupancy was evaluated at the state-level only. This data was also provided by SESAB throughout the period of March 6^th^ to May 4^th^, 2020.

We also obtained data on patients admitted to a reference infectious disease hospital located in Salvador (Instituto Couto Maia; ICM). Individual-level trajectories of 231 patients from admission to discharge/death were followed from March 23^th^ to April 16^th^. The ICM is the leading public hospital in the state of Bahia for treatment of COVID-19 patients. The collected data, aggregated across patients, informed the hospitalization-related model parameters, including the search intervals for the optimization procedures.

The estimated population of Bahia in 2020 was obtained by the Brazilian Institute of Geography and Statistics (IBGE).

### Assumptions and model construction

In this section we present the SEIIHURD model that subdivides the population into eight compartments, as follows: susceptible (*S*), those who are not exposed to the disease; exposed (*E*), individuals who have been exposed to the virus and are in a latent, non-transmittable period; (*I*) infectious, those currently infected and capable of transmitting the disease to whom they contact; recovered (*R*), those who were previously infected and either recovered from the disease or were removed as a result of death due to COVID-19 and consequently do not affect the transmission dynamics. The infectious individuals are further separated into asymptomatic, denoted *I_a_*, and symptomatic, denoted by *I_s_*. Current evidence suggests that the majority of COVID-19 infections result in mild to moderate symptoms that do not require hospitalization. However, a proportion of the infected will present with severe symptoms resulting into hospitalization (clinical beds) (*H*), while those in critical conditions will eventually require ICU admission (*U*). For simplicity, we have neglected the transmission of individuals in compartments *H* and *U*, assuming that hospital containment decreases the chances of contact with susceptible individuals. We also consider a flux of patients between the *H* and *U* compartments, as individuals initially admitted to a clinical ward may worsen their condition and require an ICU bed. Furthermore, all patients in *U* are transferred to *H* prior to discharge and recovery. In Figure 1 we present the flow diagram of the proposed model. Similar works can be found in^9,13,20,21^.

**Figure 1.**
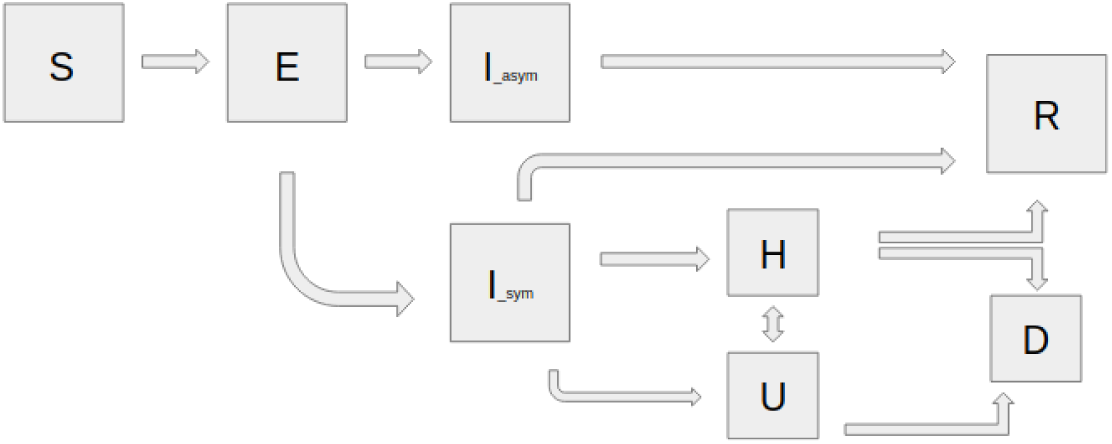
Flow diagram for modelling the dynamics of COVID-19 transmission in the 8-compartment SEIIHURD model

To account for local interventions of movement restriction (such as governmental stay-at-home orders), we consider the transmission rate as a function of time, varying according to local measures. To define *β*, let {*t*_1_, *t*_2_,…, *t_n_*} be a set of points in time defining the change on the transmission rate. Then, we can write *β* as a function of time *t* as:

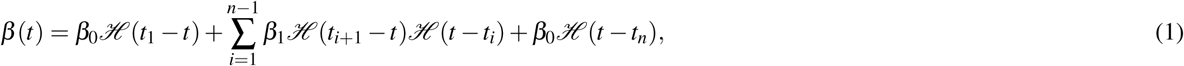

where 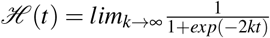 is a Heaviside step function, *β_i_* are transmission rates that can be obtained by the fitting of the data to the time interval defined by the *t_i_*’s. The system of differential equations then reads:

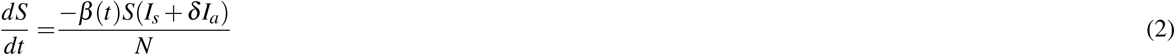

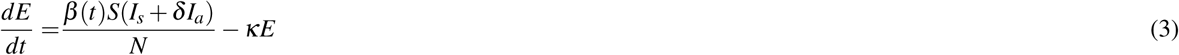

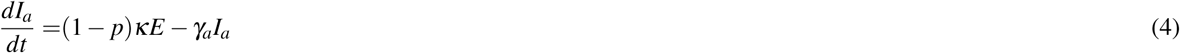

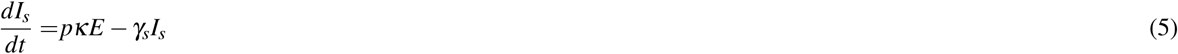

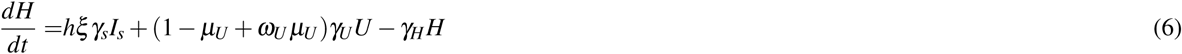

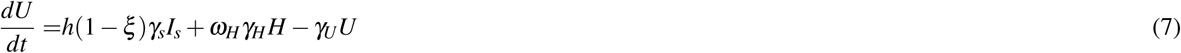

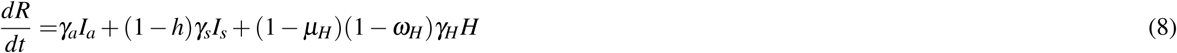

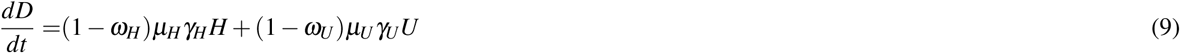

For more details about the system of equations, see Supplementary File 1.

### Analytical evaluation of ℛ_0_ and ℛ(*t*) of the SEIIHURD model

The basic reproductive number ℛ_0_ has been derived within the general next generation operator framework^21,22^. It considers the unstable disease-free equilibrium point of the model, where *S* corresponds to the whole population and all other compartments are identically set to 0. Following^22^, the value of ℛ_0_ results from a balance between the infectious and transition terms of the sub-model composed by the variables (*E*, *I_a_, I_s_*) associated with the transmission of the disease, which are gathered in the corresponding 3 × 3 matrices 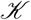 and 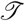, given by:

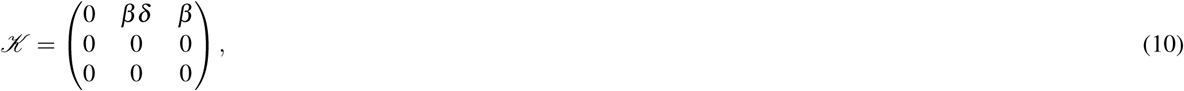

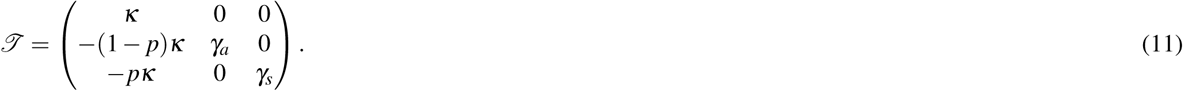

Thus, given the above matrices, ℛ_0_ corresponds to the largest eigenvalue of the matrix 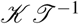 and represents the sum of the contribution of the symptomatic and asymptomatic transmission, being expressed by

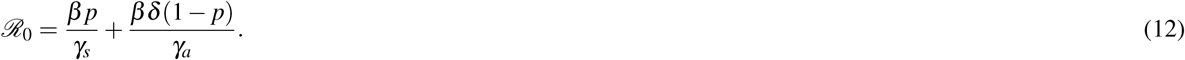

The effective reproduction number ℛ(*t*) provides a measure of how the newly infected part of the population will further transmit the pathogen as the epidemic evolves over time. Indeed, as reminded above, the evaluation of ℛ_0_ considers that the whole population is initially susceptible, a condition that is strictly valid only when the pathogen is first introduced into the system. As time evolves, the susceptible fraction of the population always decreases with time for models where the recovered compartment does not feed the susceptible one. It is still unknown whether re-infection by SARS-CoV-2 can occur, but initial evidence suggests against this possibility^23^. Here we assume that only a single COVID-19 infection event can occur in any single person.

The epidemiological meaning of ℛ(*t*) is the same as for ℛ_0_, namely, it represents the average number of secondary infections that an individual, who became infected at time t, is able to generate. The series of ℛ(*t*) values indicates the current trend of the epidemic and captures changes caused by recently-introduced interventions (such as governmental policies) or by natural decrease of the susceptible population. Accordingly, it provides a quantitative evidence of whether further measures are needed to control the epidemic. Here ℛ(*t*) has been estimated following the assumptions introduced in^24^. It is based on the series of daily infected individuals, that is considered here as the source to insert into the general renewal equation for a birth process

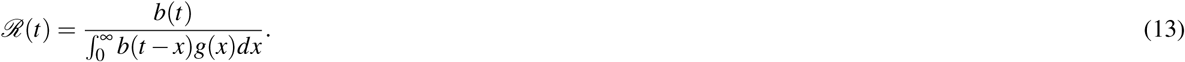

In an epidemiological context, *b*(*t*) represents the daily number of new cases, while *g*(*x*) is the disease probability distribution function for the time for an individual to infect secondary cases. In this case, the function *g*(*x*) is a contribution of the three compartments *E*, *I_a_*, *I_s_* that impact the evaluation of ℛ_0_. Therefore, we can write

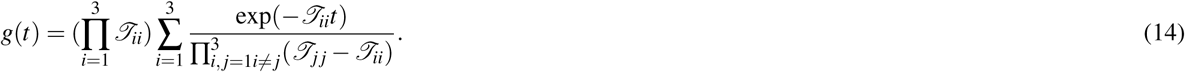

In order to overcome the fluctuations of the confirmed number of cases (which is influenced by testing capacity and its associated increase, even if momentarily, such as when pending tests accumulate), we present two series of *R_t_*, one calculated from the daily number of confirmed cases, as informed by local health authorities; and another *R_t_* series where this data is informed by the predictions of the model.

The estimation occurred within a chosen range based on literature and data collected locally, as described in Supplementary Table 1. The initial conditions (*S*_0_; *I_a,_*_0_; *I_a,_*_0_;*E*_0_;*R*_0_;*H*_0_;*U*_0_;*D*_0_) is given by (1−*I_a,_*_0_−*I_s_*_;0_−*E*_0_; *I_s_*_;0_; *I_a,_*_0_; *I_s_*_;0_;*E*_0_;0;0;0;0).

### Parameter sensitivity analysis

Sensitivity analysis was conducted to assess the effects of model parameters in the dynamics of *I_a_*, *I_s_*, *U*, *H* and *D* over time. By using an statistical variance-based method, described by Sobol^25^, the sensitivity analysis of the system described by Eqs. (2)-(9), considers the following parameter vector

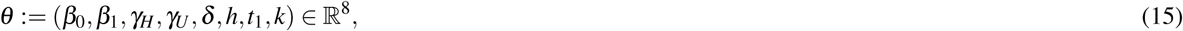

and assumes that its elements are uniformly distributed in proper intervals as follows:

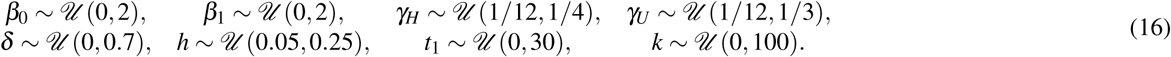

This method is divided in two steps, described in detail in Supplementary File 2. The numerical simulations were performed using the SALib library^26^, and the experiments were conducted generating *N* = 12000 samples.

### Evaluation and estimation of model parameters

The parameters *p*; *κ*; *γ_a_*; *γ_s_*; *ξ*;*ω_U_*;*ω_H_*; *μ_U_*; *μ_H_* were kept fixed and the remaining were obtained by estimating the best values that fit the model to the data (see Supplementary Table 1). To define the fixed parameters and the search intervals to use for the estimations, we performed a literature review of published paper and collected the information regarding to key epidemiological parameters that inform our model (see Supplementary File 3).

As an additional guide to obtain a locally-informed model, hospitalization data from a reference infectious disease hospital (ICM; see Data Sources section) were used and helped to evaluate the parameters. Routine clinical data were extracted to obtain estimates of mean hospitalization period (*γ_H_*), mean ICU period (*γ_U_*), death rate of hospitalized individuals (*μ_H_*), death rate of ICU individuals (*μ_U_*), proportion of clinically hospitalized transferred to ICU (*ω_H_*) and proportion of ICU individuals that return to clinical hospitalization (*ω_u_*) (see Supplementary Table 2).

The parameters of the model were estimated using the Particle Swarm Optimization (PSO) metaheuristic^27^. PSO is a population stochastic optimization technique for non-linear functions, based on the collective intelligence behavior of animals, with broad applications in the literature. Under the PSO framework, we used a multi-optimization function to simultaneously optimize the model to the series of cumulative confirmed cases and the series of cumulative deaths in the state, its capital city Salvador and the remaining inland cities up to May 4th. PSO was implemented using pyswarms library version 1.1.0 for Python 3 (http://python.org)^28^, and was executed with 300 particles through 1,000 iterations with cognitive parameter 0.1, social parameter 0.3, inertia parameter 0.9, evaluating five closest neighbors through Euclidean (or L2) distance metric. In addition to the point estimates obtained by the PSO method for the parameters *β*_0_, *β*_1_, *δ*, *h*, *γ_H_* and *γ_U_*, percentile confidence intervals were also built for these parameters. The intervals were constructed using the weighted non-parametric bootstrap method^29^, considering 500 replicates of the original series *St*, *t* = 1, …, *n*, which represents the number of new cases observed at time *t*. The proportion of observed cases at time *t* (number of cases at time *t*/total number of cases in the analyzed period) was used as a weight in the re-sampling process to obtain the bootstrap replicates. Then, the SEIIHURD model was adjusted for each replicated series and the estimates obtained for the model parameters were stored in vectors, generating the empirical distribution of each parameter.

### Modelling scenarios

We present our analysis as follows: First, we study the effect of previously enforced interventions in the state of Bahia, its capital city Salvador and the remaining inland cities. For this, we considered the SEIIHURD model with the proportion of symptomatic needing hospitalization or ICU (parameter *h* equals to zero), so that the resulting model does not consider the compartments of hospitalization, ICU and death, effectively resembling an SEIR model with asymptomatic transmission.

Second, we analyse these effects on the number of death and hospitalization requirements considering the state as a whole. Then, we show the impact of the asymptomatic infected individuals on the dynamic of COVID-19 transmission.

Lastly, to study future behaviour of the transmission of the disease in Bahia, we simulated different scenarios that may impact on the number of cases, mortality and health care infrastructure. The following scenarios were considered: (1) An immediate intervention on the May 5^th^, sustained for a period of 7, 14 or 30 days, and reducing the rate of transmission by 25%, 50% or 75%; (2) A critical intervention, adopted when the collapse of clinical bed occupancy occurs (on 05/14/20, the predicted date), maintained for a period of 7, 14, 30, 60, 90 days and reducing the rate of transmission by 25%, 50% or 75%; (3) and last, implementations of more than two interventions in sequence.

### Data availability

Codes used to produce the results presented herein, and related datasets, are available in a public GitHub repository^30^.

### Ethics statement

This study was conducted with anonymized secondary data, where only aggregated clinical data was used for model parameterization. Therefore, no approval from a Human Research Ethics Committee is required.

## Results

### Effects of social distancing and governmental interventions in disease transmissibility can be observed shortly after onset

We started our analyses by assessing the effects in disease transmissibility that local non-pharmaceutical interventions (NPI) have produced in the state, in its capital, as well as in the inland cities (all 417 municipalities in the state except the city of Salvador). For this, the model was fitted using the number of confirmed cases obtained by local authorities (Figure 2) and we estimated parameters related to the transmission rate (*β*_0_, *β*_1_), the timepoint in their change, and the factor that reduce the asymptomatic infectivity *δ*. It can be observed that the initial (pre-intervention) transmission rate was *β*_0_ = 1:06 ([1.04 −1.07] 95% CI). A reduction of 38% on the transmission rate, yielding *β*_1_ = 0:63 ([0.62, 0.65] 95% CI), is evidenced around March 30th (24 days after the first confirmed case in the state). In Salvador, the capital of the state (with a population of 2.6-million people), this reduction was of 52.4% (on March, 26th), while inland cities displayed a decrease of approximately 34% (on April, 1st). The factor that reduces the infectivity of the asymptomatic was determined to be *δ* = 0:62 ([0.60, 0.64] 95% CI) at the state-level, *δ* = 0:55 ([0.54, 0.56] 95% CI) in the capital and *δ* = 0:57 ([0.56, 0.58] 95% CI) in the inland cities. The basic reproduction numbers for Bahia, the capital and inland cities were respectively, of *R*_0_ = 2:7 ([2.6 −2.8] 95% CI), 2:9 ([2.8 - 3.0] 95% CI) and 2:3 ([2.2 - 2.4] 95% CI).

These results show that the combined effects of changes in human behavior with governmental policies of movement restriction result in significant decreases in the transmissibility of the disease, as measured by the *β* parameter. However, these efforts are insufficient to curb the epidemic in the state, as the basic reproductive number exceeds 1, indicating a scenario of continuing growth of the epidemic.

### Projecting hospitalization requirements in Bahia, Brazil: challenges for low-resource settings

Next, we evaluated the burden on hospitalization needs imposed by the COVID-19 epidemic at the state-level, as well as the effects that NPI strategies could have on these requirements. We also estimated the total number of deaths projected by the model in the absence and with the enforcement of distancing measures. Our results, presented in Figure 3 show that, in the absence of interventions, the state-level availability for clinical beds would be exhausted by April 22^nd^. With the maintenance of the current level of interventions, this depletion is shifted in time and would occur by May 14^th^. Analogously, the demand for ICU beds would exceed their capacity by April 23^rd^ in the absence of interventions, and by May 17^th^ with the current rate of interventions.

The real-world, state-level data obtained for May 4^th^, the last day with available bed occupancy observations, shows that 240 (51.5%) clinical beds were occupied, while 176 (41.7%) ICU beds were in use. Our model-based analysis yields an increase in these numbers by 29 (7,153 places) and 31 (5,517 places) times if interventions had not been adopted in the state. On the other hand, the enforcement of measures decreased the number of cases and deaths by 20 and 14 times, respectively.

These results underscore the impact that the ongoing COVID-19 epidemic imposes on hospital resources, and eventually on deaths, and particularly explicits the challenges faced by countries with lesser-developed healthcare systems. Even if we consider overestimation of the prediction results, the simple doubling of the current real-world bed occupancy data would already result in exceeding the current availability of clinical and ICU beds in the state.

**Figure 2.**
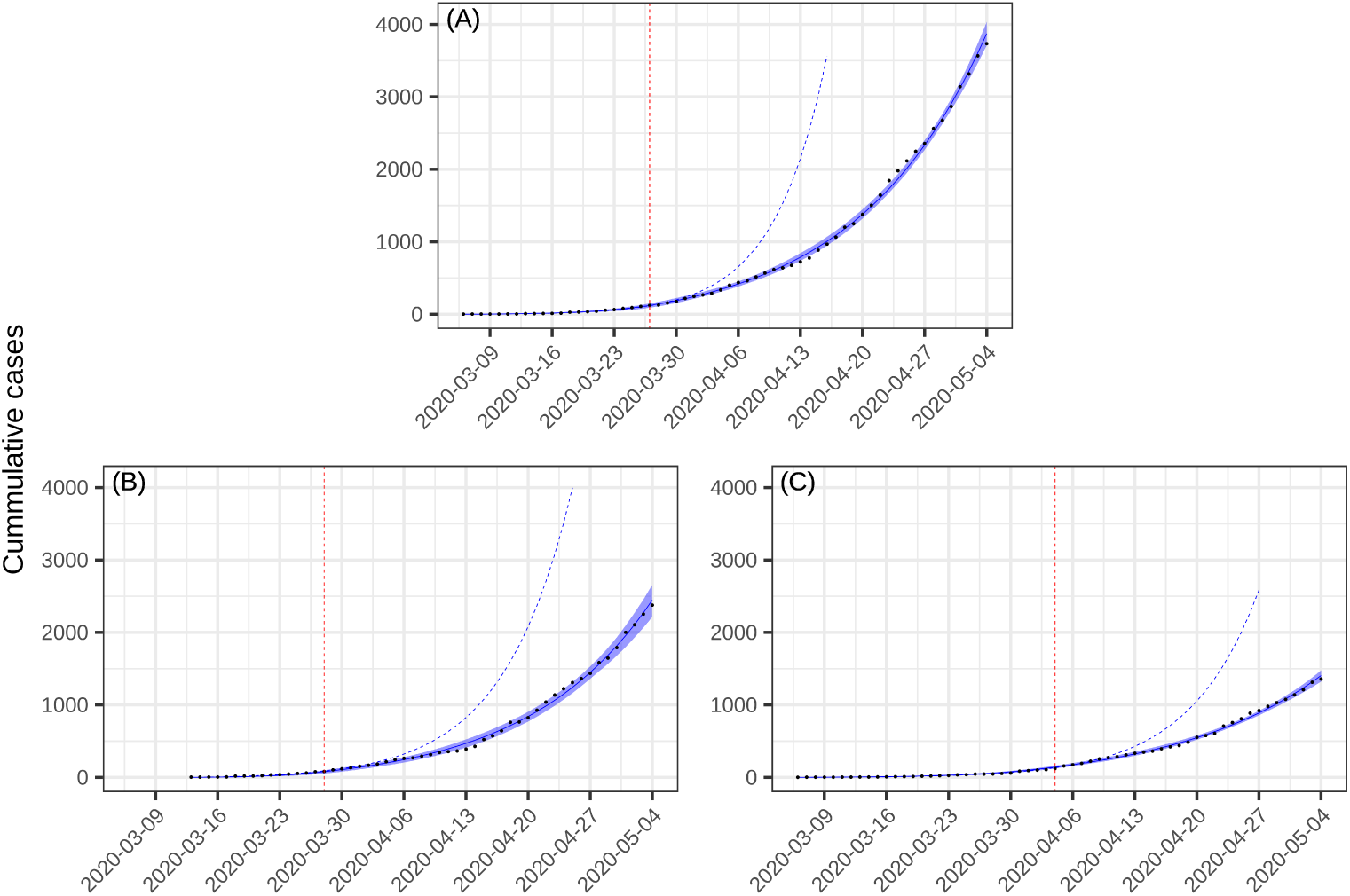
Projection of the the number of cases with a changing transmission rate (a) in Bahia; (b) in Salvador and (c) in the inland cities. The parameters *κ* = 1/4, *p* = 0:2, *γ_a_* = 1=3:5, *γ_s_* = 1=4 were fixed and h set to zero. The black dots correspond to the actual number of cases. The vertical dashed red lines are the dates of transition from *β*_0_ to *β*_1_. The blue dashed and full lines represent the evolution of the epidemic with a fixed transmission rate *β*_0_ and with both *β*_0_ and *β*_1_, respectively.

The obtained ℛ_0_ is as before and the effective reproductive number is presented in Figure 4. We can notice a trend of reduction on the effective reproductive number, although ℛ(*t*) is above one in almost the complete time series for the state of Bahia. Interestingly, our analysis of how asymptomatic individuals influence the course and dynamic of transmission revealed that these individuals contribute to an increase of 68.62% on the basic reproduction number.

### A model-informed strategy of periodic interventions to reduce COVID-19 transmissibility as a cue to protect health systems

The previous results revealed the favorable effects of interventions that result in decreases in the disease transmission rate on shifting the peak of hospitalization saturation (complete occupancy of available beds), and in decreasing the number of cases and deaths. However, these results showed that complete saturation is inevitable under our local conditions. We next evaluated to what extent more vigorous restriction policies, and their duration/periodicity, would be useful in order to prevent the complete collapse of the state-level health system. To address this question, we used the described model to study the epidemic dynamics under various scenarios.

Initially, and in order to assert that disease transmissibility is the driving factor leading to increased hospitalization requirements, we considered a scenario where the transmission rate of asymptomatical individuals was increased by 50% starting on May 5^th^ (Figure 5). After 20 days, we noticed an increase of cumulative cases and deaths of, respectively, 50%, and 37%. Accordingly, clinical and ICU bed requirements increase by 114% and 100%, respectively. This scenario would be illustrative of a situation where the movement restriction of asymptomatic individuals is eased.

The previous results confirmed the importance of curbing disease transmissibility. Then, we turned to set targets that would allow for an increase in the protection of the health care system. These scenarios are illustrated in Table 1. We show that an intervention that reduces the transmission rate to 25%, enforced on May 5^th^ for 14 days, would not yield significant improvements, resulting in a gain of only 3 days until clinical beds collapse, and 10 days until ICU bed capacity is exhausted. Similar results can be achieved by a more punctual (7 days period), but more vigorous intervention reducing transmission rate by 50% (Table 1). More interestingly, a delay of about 40 days for clinical and ICU bed exhaustion can be achieved in a scenario where a 50% reduction on the transmission is sustained for 30 days, or when a 75% reduction is endured for 14 days (Supplementary Figure 1).

**Figure 3.**
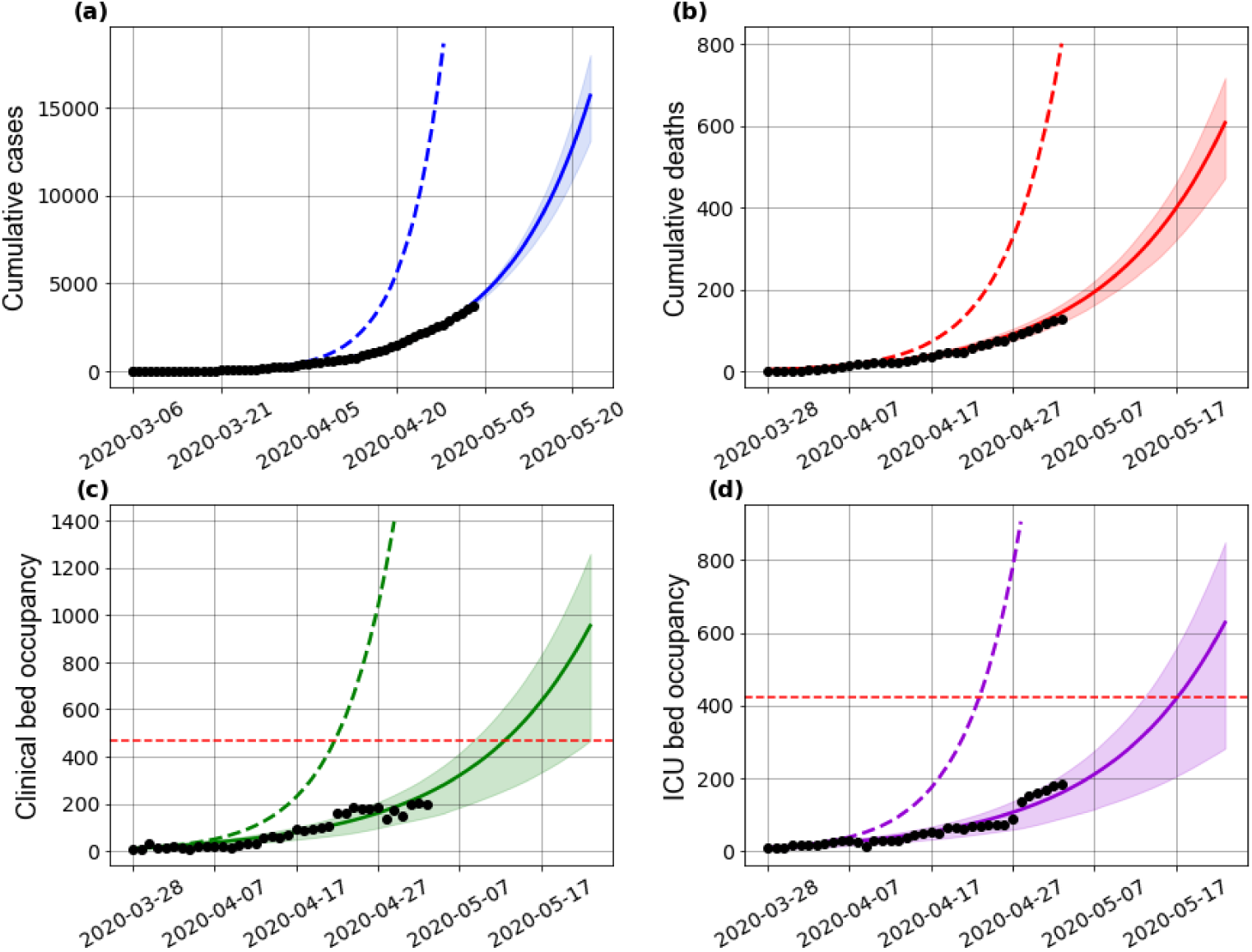
Effects of the implemented interventions on the (a) number of cases, (b) deaths, (c) clinical hospitalization and (d) ICU bed requirements at the state-level. The horizontal red dashed lines are, respectively, the current capacity for beds for clinical hospitalization (466 beds) and ICUs (422 beds). The non blue dashed and full lines represent the evolution of the epidemic with a fixed transmission rate *β*_0_ and with both *β*_0_ and *β*_1_, respectively. The assumed parameter values are shown in Supplementary Table 1.

**Table 1.** Scenarios of an immediate intervention on May 5th in Bahia, with variations of the transmission rate and intervention length.

| Percentage of transmission rate reduction | Intervention length (days) | Date of hospitalization beds collapse (delay, in days, compared to baseline scenario) | Date of ICU beds collapse (delay, in days, compared to baseline scenario) |
| --- | --- | --- | --- |
| 25% | 7 | 05/17/20 (3) | 05/21/20 (4) |
|  | 14 | 05/18/20 (4) | 05/24/20 (7) |
|  | 30 | 05/18/20 (4) | 05/27/20 (10) |
| 50% | 7 | 05/21/20 (7) | 05/26/20 (9) |
|  | 14 | 05/31/20 (17) | 06/04/20 (18) |
|  | 30 | 06/22/20 (39) | 06/26/20 (40) |
| 75% | 7 | 05/27/20 (13) | 06/01/20 (15) |
|  | 14 | 06/13/20 (30) | 06/17/20 (31) |
|  | 30 | 07/21/20 (68) | 07/24/20 (68) |

The timing of interventions is also crucial under our model. If a vigorous intervention is only adopted on the day when clinical bed occupancy reaches its maximum availability (on May 14^th^), a time lag will be needed in order to allow for patient turnover in hospitals. Once occupancy is below the total availability, interventions can be continued or suspended. In the latter case, hospitalization requirements resumes its growth until reaching the health system’s capacity once again. Under this scenario, an intervention that reduces the transmission rate by 25% will not be enough to protect health resources, even if policies are maintained for long periods of time (Figure). Similar results are seen when we consider the reduction of the transmission rate by 50%, as shown in Table 2. Thus, harsher efforts to contain disease transmissibility, and for more extended periods, are necessary to allow for full recovery of the healthcare system.

**Figure 4.**
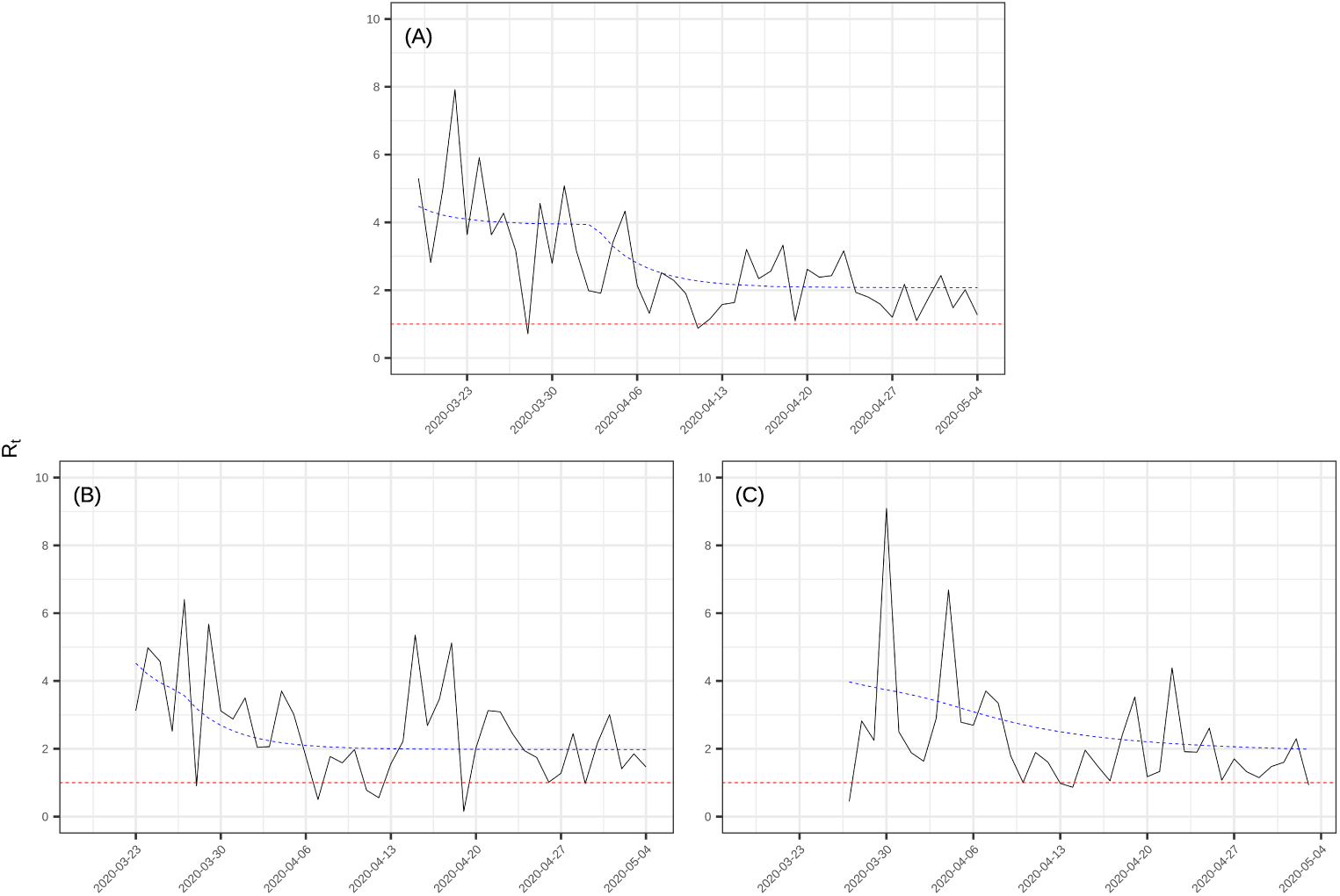
Effective reproductive number, given by Equation (13) for (a) Bahia, (b) Salvador and (c) inland cities up to May 4^th^, 2020. Black line represents the *ℛ_t_* calculated with reported number of new cases; the blue dashed line represents the *ℛ_t_* calculated with the new number of simulated cases obtained from the model. The black dots correspond to the actual number of cases (a), deaths (b), and hospital bed occupancy (c)-(d).

**Figure 5.**
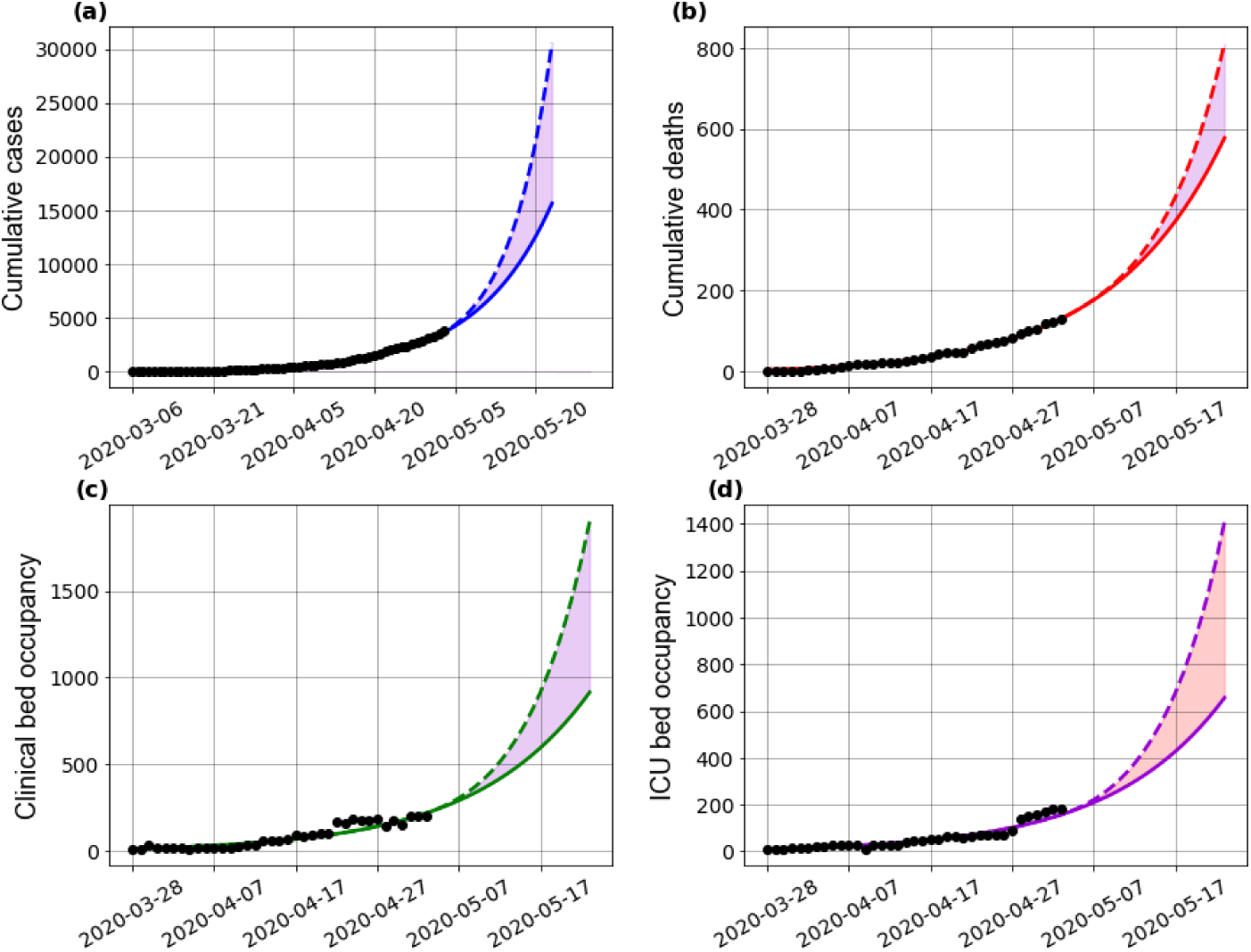
Effect of easing the social distancing for asymptomatic individuals in Bahia on the (a) number of cases, (b) deaths, (c) clinical hospitalization and (d) ICU bed requirements at the state-level. Here, the transmission rate of asymptomatic cases has been increased by 50%, by setting *δ =* 0.93. The black dots correspond to the actual number of cases (a), deaths (b), and hospital bed occupancy (c)-(d). The assumed parameter values are shown in Supplementary Table 1.

**Table 2.** Scenarios of critical interventions adopted exactly when total clinical beds availability is reached (05/14/20), with variations of the transmission rate and intervention length

| Percentage of transmission rate reduction | Intervention length (days) | Date when the hospitalization threshold is re-achieved (days) | Date of second hospitalization collapse (days of delay) | Date when the ICU threshold is re-achieved (days) | Date of second ICU collapse (days of delay) |
| --- | --- | --- | --- | --- | --- |
| 50% | 7 | does not occur* | — | does not occur* | — |
|  | 14 | does not occur* | — | does not occur* | — |
|  | 30 | does not occur* | — | 06/05/20 (19) | 06/25/20 (20) |
|  | 60 | 06/23/20 (40) | 08/03/20 (41) | 06/06/20 (19) | 08/06/20 (62) |
|  | 90 | 06/23/20 (40) | 09/14/20 (83) | 06/05/20 (19) | 09/17/20 (104) |
| 75% | 7 | does not occur* | — | 05/27/20 (9) | — |
|  | 14 | 05/31/20 (17) | 06/08/20 (8) | 05/27/20 (9) | 06/16/20 (30) |
|  | 30 | 05/31/20 (17) | 07/21/20 (51) | 05/27/20 (9) | 07/24/20 (68) |
|  | 60 | 05/31/20 (17) | 09/29/20 (121) | 05/27/20 (9) | 10/02/20 (138) |

\*, hospitalization is not reduced enough to reach the actual capacity.
—, does not apply.

The previous results, combined, show that intense efforts to decrease COVID-19 transmissibility are needed in order to overcome a complete collapse of the health care system in a low-resource setting such as that encountered in Bahia, Brazil. Of note, under some of the presented scenarios full establishment of the hospitalization capacity may not be achieved if the timing to enforce more strict measures to decrease the transmission rate is not optimal. Accordingly, periodic interventions may be needed to control secondary waves of the outbreak. In Figure 6 we illustrate the behaviour of the spread dynamics of COVID-19 in Bahia if measures are periodically adopted.

### Model sensitivity analysis

A sensitivity analysis was performed in order to evaluate the most influential parameters of the model (Figure 1 within Supplementary File 2). These results revealed that the factor that reduce the asymptomatic infectivity, *δ*, is among the most influential parameters to every model output during the whole period evaluated. Also, the transmission rate *β* was identified as exerting an important role for model output, as expected from our previous analyses. Particularly, during the first 30 days *β*_0_ is the most important parameter in the system, as indicated by higher values of S_T_ (Figure 1 within Supplementary File 2). After this period, the importance of *β*_0_ decreases as that of *β*_1_ increases, eventually superseding the former as the most important parameter in the system. For H, U, and D, the most influential parameter during the initial stages of the simulation (before day 15) is the proportion of symptomatic needing hospitalization or ICU, *h*.

## Discussion

The COVID-19 pandemic is posing unprecedented challenges in healthcare resources worldwide. The results presented in this work, based on actual epidemics data and on a generalization of the SEIR model taking into account asymptomatic cases, hospitalization demands and mortality, make evident some relevant scenarios for COVID-19 in Bahia, a Brazilian state with strong inequalities in health coverage and access. The epidemic trajectory can be well characterized by the basic reproductive number (ℛ_0_ > 1), indicating the exponential growth of cases at the beginning of the epidemic in Bahia, its capital city Salvador and well as the 416 inland municipalities. We showed that a reduction on the disease transmission rate, as a result of non-pharmacological governmental interventions initiated on March 17^th^, led to a decrease in the number of cases, hospitalization demands and mortality up to May 4^th^, which is represented in the model by using a step function of the transmission rate. We show a reduction of 38% of the transmission rate during 2 months since the first confirmed case in the state of Bahia. This may be partly attributed to population adherence to social distancing recommendations and convergent action of local government authorities at the state and the municipalities levels. The effect of social distancing is also apparent on the time series of the effective reproduction number, ℛ(*t*), in spite of the number of secondary cases generated by the previous cases continuing to be greater than 1 (ℛ(*t*) > 1) in this ongoing epidemic.

Several measures to control the spread of the disease have been enforced by the local government, some of them even before the notification of the first cases of community spread, on March 19^th^. From 17^th^–28^th^ March, measures were gradually applied, and included the suspension of public gatherings of over 50 people, closure of schools, mandatory home isolation for people with respiratory symptoms, adoption of teleworking for workers belonging to risk groups, and the reduction of circulation of interstate buses and intercity transportation from places where SARS-CoV-2 community transmission was declared. Concerning the latter, it is possible that the transmissibility decrease observed for the capital led to a corresponding reduction in inland cities due to decreased transportation flux of individuals.

**Figure 6.**
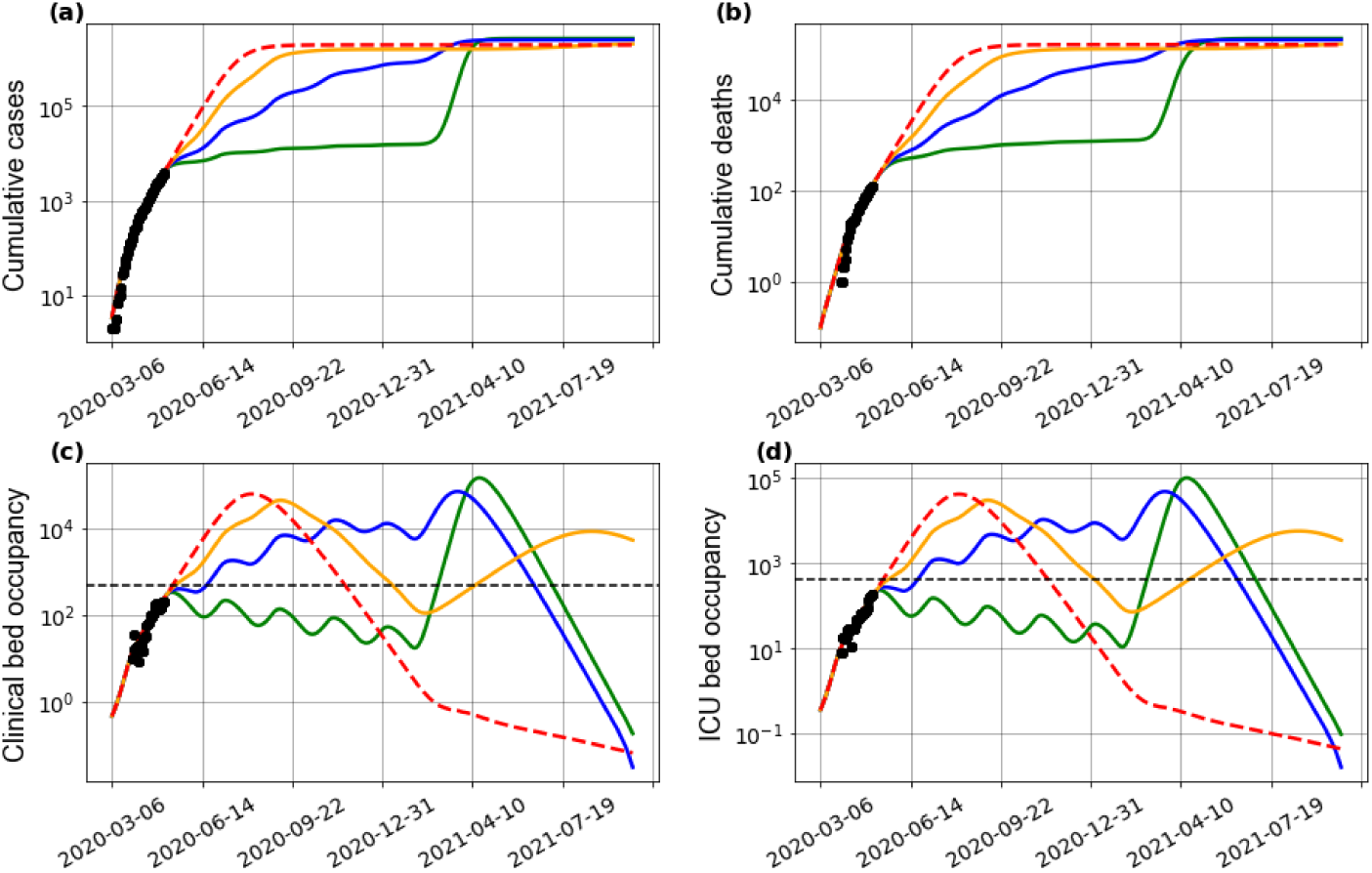
Effect of periodic interventions on the (a) number of cases, (b) deaths, (c) clinical hospitalization and (d) ICU bed requirements at the state-level. The transmission function *β*, as in Eq. 1, is defined by considering a reduction of 25% (yellow curves), 50% (blue curves) and 75% (green curves) on the *β*_1_ parameter. The red curves considers a scenario of no reduction in *β*_1_, and the period is the intervention window applied. The dashed horizontal lines in (c) and (d) indicate the total number of clinical and ICU beds available in the state, respectively. The black dots correspond to the actual number of cases (a), deaths (b), and hospital bed occupancy (c)-(d). The used parameter values are in Supplementary Table 1.

Given the extent of measures adopted from March 17^th^ onward, our results show that the reduction of the demand for clinical and ICU beds possibly avoided a surge in hospitalization needs leading to immediate system collapse, at least up to May 15^th^. However, the scenarios simulated revealed that easing social distancing measures abruptly, leading to increased transmission rates, should not be considered due to the non-linear transmission of the disease and the significant number of asymptomatic infections, which have been considered as the source for the majority of cases in previous studies^9^. Our results reinforce the negative effects on healthcare resources related to the circulation of asymptomatic cases. Accordingly, policies aiming at easing the current level of social distancing measures, in a scenario where the majority of the population does not have access to tests, could pose an additional burden on an already limited health system infrastructure. These results are in line with a recent study suggesting self-isolation of all individuals, irrespective of symptoms, as a strategy to cope with the increasing demand for COVID-19-related hospitalization^17^.

The consequences of the spread of the disease are even worse when the healthcare system is no longer able to support the amount of patients needing specialized assistance–a situation declared as a collapse. Our results point that non-pharmaceutical measures should be implemented in order to reduce the transmission rate of the disease, and consequently gain time to create new hospitals, acquire protective equipment material and guarantee human resources. But what can be done when faced with an already collapsed health system? We performed simulations to address this question, presenting different scenarios in order to determine an efficient strategy, by considering the period and intensity of interventions. The interventions can be applied at a single moment in time and kept until a decrease of the number of cases is observed, or a combination of intervention can be enforced at different time intervals^31,32^, depending on testing and monitoring capacities and/or local social-economical conditions. Our results show that when faced with an already collapsed system, only vigorous measures (that reduce the transmission rate by at least 50%) enforced over at least two months, or alternatively measures capable of reducing transmission by at least 75%, over a 2-week period, are capable of re-establishing hospitalization operation capacity. Albeit harsh, other countries have successfully managed to reduce transmission at such figures by employing a myriad of public health measures (including intra-city and inter-city travel restrictions, social distancing, home confinement and centralized quarantine and expansion of available medical resources)^33^. Of note, even in the event that transmission rate is decreased at these levels, a further, second collapse on hospitalization needs cannot be completely averted.

Our findings have some limitations. First, this study was carried out with routine local data, which may result in an underestimation of the real incidence of COVID-19, an avoidable problem also detected in other diseases^34^. Mass testing is still not performed in the country, and current policies recommend that individuals suspected of COVID-19 infection should only seek health care assistance when presenting with mild-to-moderate symptoms. However, the current available national surveillance data can be considered adequate for the identification of trends of the disease, as this system is standardized and implemented in all municipalities in the country. Nevertheless, we were able to parametrize our model to a more realistic setting by using hospitalization data from a local reference infectious disease hospital currently dedicated to the care of COVID-19 patients, and the results were compared to model parameters obtained from the state of Bahia as well as the literature. Thus, our modelling strategy has as an advantage being locally-informed, yielding more realistic results. The implemented model does not consider transmission of infected individuals undergoing hospitalization, although it is known that health care workers are more at-risk of many airborne infections, and transmission is particularly high during procedures that generate aerosols^35^. In spite of these limitations, given the general character of the mathematical model described herein, it may be readily applied to other places current tackling the COVID-19 epidemic with simple re-estimation of the assumed values for parameters, and by taking into account the local COVID-19 epidemic panorama.

By drawing on different modelling scenarios, in this work we determined whether an efficient strategy can be employed to avoid a collapse of the health care system, taking into account the length and the intensity of interventions. There exists a compromise between the availability of hospital beds and the pool of susceptible individuals, which eventually leads to a second wave of epidemics, and further shortages of hospital beds. Our results underscore the importance of carefully consideration of the suspension of restrictive measures by policy-makers.

## Data Availability

The data that support the findings of this study are openly available in Mathematical and Statistical Modeling of COVID-19 in Brazil at https://github.com/cidacslab/Mathematical-and-Statistical-Modeling-of-COVID19-in-Brazil.

## Acknowledgements

This study was financed in part by the Coordenação de Aperfeiçoamento de Pessoal de Nível Superior – Brazil (CAPES) – Finance Code 001. STRP was supported by an International Cooperation grant (process number INT0002/2016) from Bahia Research Foundation (FAPESB). STRP and RFSA were supported by the National Institute of Science and Technology - Complex Systems from CNPq, Brazil. JFO was supported by the Center of Data and Knowledge Integration for Health (CIDACS) through the Zika Platform- a long-term surveillance platform for the Zika virus and microcephaly, Unified Health System (SUS) - Brazilian Ministry of Health. The authors acknowledge the CoVida Network.

## Additional information

**Supplementary File 1. Differential Equations extended**

**Supplementary File 2. Parameter sensitivity analysis**

**Supplementary File 2. Key epidemiological parameters used in the SEIIHURD model, with their mean estimates (or range[s]) obtained from the literature**.

**Supplementary Table 1. Key epidemiological parameters used in the SEIIHURD model, with their mean estimates and respective estimates obtained according to best fit for Salvador, inland cities and all Bahia state. The parameters interval was consolidated by Supplementary Table 2 and File 3**.

**Supplementary Table 2. Epidemiological parameters associated to hospitalization dynamics of patients with Covid-19 from the Couto Maia Institut**.

**Supplementary Figure 1** Effect of the enforced interventions on the number of deaths, clinical hospitalization and ICU requirements in Bahia after immediate intervention on the 5th of May, under the scenarios of 25%, 50% and 75% reduction of the transmission rate *β*_1_. The different colours represent the period of maintenance of the measures as: 7, 14, 30, 60 and 90 days. The red dashed line represent the scenario where no change in the transmission rate is attained.

**Supplementary Figure 2** Effect of the enforced interventions on the number of deaths, clinical hospitalization and ICU requirements in Bahia on May 14th and reduction of 25% of the transmission rate *β*_1_. The different colours represent the period of maintenance of the measures as: 7, 14, 30, 60 and 90 days. The blue line represents the current scenario.

